# Reduced fecal intestinal alkaline phosphatase is associated with gestational diabetes mellitus: A hospital-based multicentre cross-sectional study in Bangladesh

**DOI:** 10.64898/2026.05.14.26353231

**Authors:** Prodipta Chowdhury, Tania Tofail, Nazia Akter, Hasanul Islam, Arnab Bokshi, Mariam Sultana, Smeeta Podder, Madhu S. Malo, MA Hasanat

## Abstract

Gestational diabetes mellitus (GDM) is a major metabolic complication of pregnancy with significant maternal and fetal adverse consequences. Beyond classical mechanisms, emerging evidence suggests that gut-derived metabolic endotoxemia may contribute to dysglycemia. Intestinal alkaline phosphatase (IAP), a key enzyme involved in maintaining gut barrier integrity and detoxifying lipopolysaccharides, has been linked to type 2 diabetes mellitus; however, its role in GDM remains largely unexplored. This hospital-based cross-sectional analytical study evaluated fecal IAP levels and their association with GDM among 198 pregnant women recruited from three antenatal care clinics representing three tiers of ANC services. Participants were screened for GDM using a 75-g oral glucose tolerance test and classified as having GDM (n=55) or normal glucose tolerance (NGT; n=143) according to WHO 2013 criteria. Stool samples were collected, and fecal IAP levels were measured using an enzymatic colorimetric assay. Fecal IAP level was significantly lower in women with GDM than in those with NGT (median 23.59 vs 46.48 U/g stool; p<0.001). Lower IAP level remained independently associated with GDM after adjustment for body mass index and previous GDM (adjusted OR 0.98 per unit increase; 95% CI 0.97–0.99; p<0.001). A graded relationship was observed between declining IAP level and GDM. Receiver operating characteristic analysis demonstrated modest discrimination (AUC 0.676), while a threshold of approximately 65 U/g stool yielded high sensitivity (89.1%) but lower specificity. Reduced fecal IAP is independently associated with GDM, supporting a potential role of gut-derived metabolic dysregulation in gestational glucose intolerance. While not suitable as a standalone diagnostic tool, fecal IAP may serve as a complementary biomarker for risk stratification during pregnancy. Prospective studies are warranted to determine its predictive value and explore its potential as a therapeutic target.

## Introduction

Gestational diabetes mellitus (GDM) is the most common metabolic complication of pregnancy [1], characterized by glucose intolerance recognized during the second or third trimester in women without pre-existing diabetes [2–4]. With global prevalence affecting 7% to 25% of pregnancies worldwide [5], GDM poses significant short- and long-term health risks to both mother and neonate, including a predisposed risk for developing Type 2 Diabetes Mellitus (T2DM) later in life [6,7].

The etiology of GDM is multifactorial, traditionally attributed to two primary mechanisms: diminished insulin sensitivity or impaired secretion, and the antagonistic effects of placental hormones—including human placental lactogen, prolactin, progesterone, and glucocorticoids, which collectively exacerbate maternal insulin resistance [8]. In GDM, these physiological shifts are pathologically exacerbated or inadequately compensated by insulin production. Given these shared defects in insulin dynamics, GDM is increasingly viewed as a specific subtype or precursor to T2DM, with researchers suggesting both conditions may represent different manifestations of the same metabolic entity [9]. Notably, while obesity is a common driver of resistance, the existence of lean GDM phenotypes suggests that a primary secretory defect and other mechanisms might play a role in the disease’s pathogenesis [10].

Intestinal alkaline phosphatase (IAP) is a membrane-bound glycoprotein and an established marker of enterocyte differentiation expressed primarily in the proximal small intestine [11,12]. As one of four human alkaline phosphatase (AP) isoforms—alongside tissue-nonspecific (TNAP), placental (PLAP), and germ-line-specific (GCAP)—IAP serum levels remain remarkably stable during pregnancy, even as PLAP and bone-derived TNAP activities rise significantly [11,13,14]. IAP is secreted bidirectionally into the lumen and circulation, maintains microbial homeostasis, limits lipid absorption, preserves mucosal integrity, and detoxifies bacterial toxins, such as lipopolysaccharides (LPS) and flagellin, via phosphohydrolysis [15–17]. IAP accounts for approximately 80% of fecal AP activity, and its reduction is a recognized feature of inflammatory bowel disease, celiac disease, and T2DM [18,19]. IAP deficiency is a potent correlate of T2DM; specifically, a large prospective study demonstrated that a 25 U/g reduction in IAP level is associated with a 35% increase in T2DM risk, an effect that remains independent of glycemic status, medication use, or obesity. In GDM, functional shifts in the gut microbiota—characterized by increased Gram-negative bacteria and lipopolysaccharide (LPS) biosynthesis—may facilitate LPS translocation into the systemic circulation [20,21], resulting in metabolic endotoxemia that triggers low-grade inflammation, impairs insulin signalling, and disrupts glucose homeostasis [22].

Despite mechanistic overlap between GDM and T2DM, the role of IAP in pregnancy remains unexplored. By addressing this gap, this study sought to quantify fecal IAP levels and determine the prevalence of IAP deficiency (IAPD) in pregnant individuals, specifically evaluating the association between IAP level and the GDM status. To our knowledge, this represents the first investigation into IAP as a potential biomarker for gestational glycemic dysfunction. We hypothesized that fecal IAP would be lower in women with GDM than in normoglycemic pregnant women.

## Methods

### Study Population

This was a hospital-based cross-sectional analytical study with a case–control comparison between women with GDM and those with normal glucose tolerance (NGT) recruited from three antenatal care (ANC) clinics representing three tiers of ANC services in Bangladesh—a rural primary care ANC clinic, a suburban medical college hospital ANC clinic, and an urban university hospital ANC clinic—and attending a gestational diabetes mellitus screening clinic between March 2024 and September 2025. Inclusion criteria comprised all singleton pregnancies screened for GDM during the study period. Women with multiple gestations, pre-existing diabetes, suffering from any acute illness (e.g. Gastroenteritis, RTI, UTI etc.), diagnosed previously with GI diseases (coeliac disease, IBD), taking steroids, anti-inflammatory agents, immunomodulators, protease inhibitors, laxatives, NSAIDs, chronic PPI user (standard dose PPI for >8 weeks) or had recently used (within 6 weeks) antibiotics were excluded. Ethical approval was obtained from the Institutional Review Board of BMU [Registration no. 4823 (BSMMU/2024/2012); date: 25/Feb/2024], and written informed consent was obtained from all participants. Confidentiality was maintained in accordance with the Declaration of Helsinki.

### Study Design

Eligible women were recruited from the participating ANC clinics of the respective Obstetrics and Gynaecology departments and were instructed to attend the corresponding facility-associated GDM screening clinics of the Endocrinology departments in a fasting state. Participants were given a stool pot on the day of recruitment & advised to attend the GDM clinic with a morning stool sample collected by themselves in a fasting state of at least 8–10 hours. Upon arrival at the GDM clinic, the investigator collected the stool pot and stored it at 4°C after proper coding and labelling. Written informed consent was taken from all patients. Demographic, family history, medical and obstetric history, anthropometrics, and other related information of each subject were recorded in a data sheet. Then, each subject was investigated using three samples of a 75-g Oral glucose tolerance test (OGTT). The fluoride tubes for glucose estimation were sent to the Department of Biochemistry, and plasma glucose was analysed on the same day of collection. If the test met the diagnostic criteria for GDM set by the WHO in 2013, they were enrolled as having GDM. Those who did not meet the GDM criteria served as controls, and those diagnosed with DIP were excluded (Fig 1). Stool samples were sent to the Stap BioTech lab in Ashulia, Dhaka, weekly, and stool alkaline phosphatase was measured there. During transport, samples were kept in a cool box and maintained at 4°C until assay. In the Stap BioTech lab in Ashulia, it has been validated that stool IAP remained stable at 4 °C for up to 15 days, and our maximum time between stool collection and assay was 07 days.

**Fig 1.**
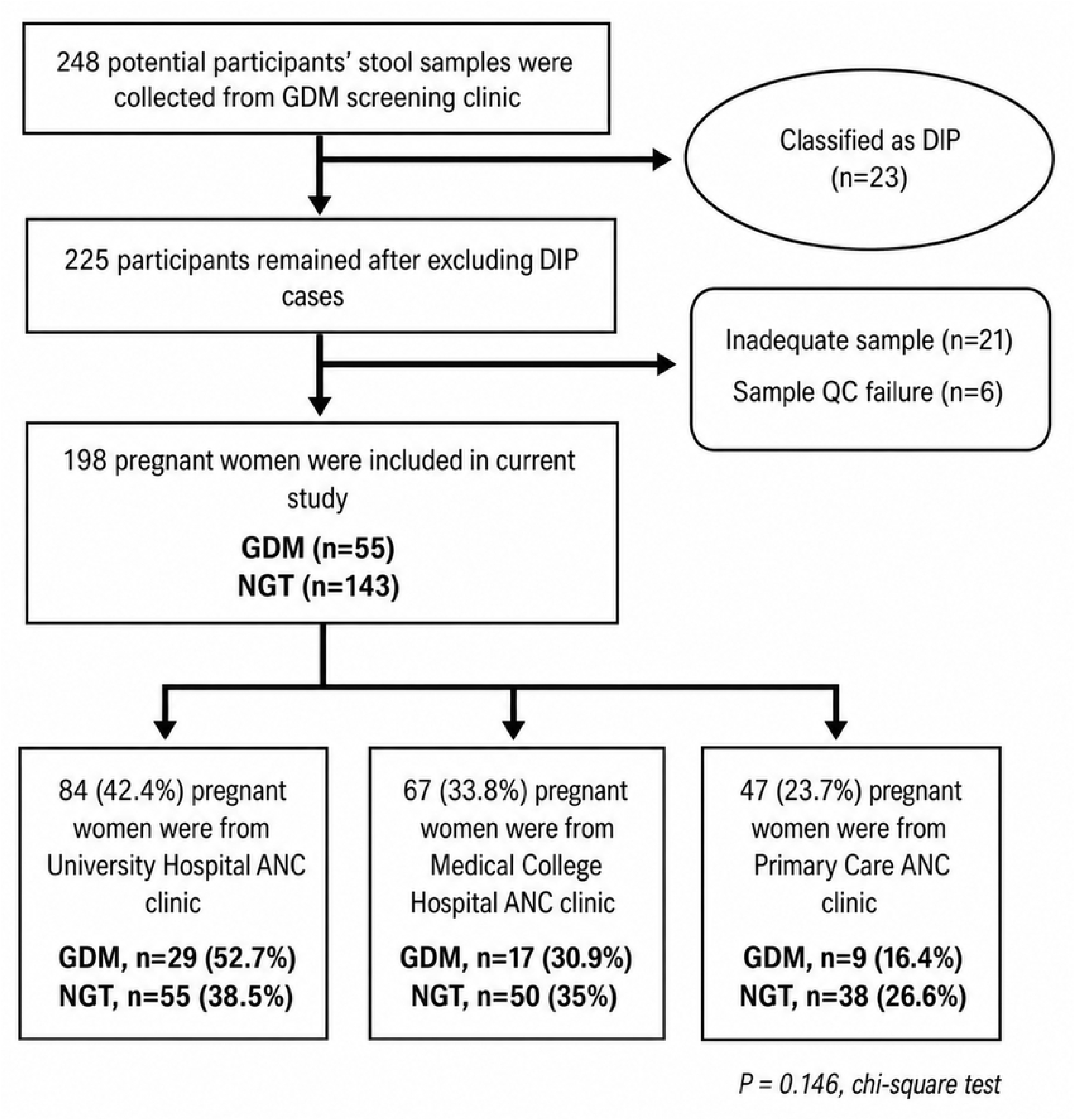
Study flowchart. Stool samples from 248 potential participants were collected at the GDM screening clinic. Of these, 23 participants were classified as having diabetes in pregnancy (DIP). Among the remaining 225 participants, 21 were excluded because of inadequate samples and 6 because of sample quality-control failure. Finally, 198 pregnant women were included in the current study: 55 with GDM and 143 with normal glucose tolerance (NGT). Among the included participants, 47 (23.7%) were from the primary care ANC clinic [GDM, n=9 (16.4%); NGT, n=38 (26.6%)], 67 (33.8%) were from the medical college hospital ANC clinic [GDM, n=17 (30.9%); NGT, n=50 (35.0%)], and 84 (42.4%) were from the university hospital ANC clinic [GDM, n=29 (52.7%); NGT, n=55 (38.5%)]. Recruitment-source distribution did not differ significantly between the GDM and NGT groups (χ^2^ test, p=0.146). Abbreviations: **ANC**, antenatal care; **DIP**, diabetes in pregnancy; **GDM**, gestational diabetes mellitus; **NGT**, normal glucose tolerance; **QC**, quality control.

### Definitions

**Gestational diabetes mellitus (GDM) and Diabetes mellitus in pregnancy (DIP)** were diagnosed by the WHO-2013 criteria

**IAPD (Intestinal ALP deficiency):** <65 U of IAP per gram of stool was considered a deficiency (IAPD) [18]

### Sample Size

Sample size was estimated for an unmatched case–control design using the Kelsey formula (equal allocation):

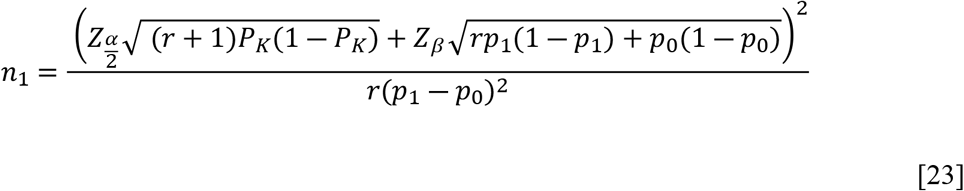

Here, Control exposure prevalence (IAPD in NGT) *p*_0_ = 0.398 [18]; Odds ratio, OR = 3.27 [24]; Case: control ratio 1:1 → *r*=1; α = 0.05 (two-sided) → Z_α/2_ = 1.96; Power = 90% → Z_β_ = 1.2815515655. Using this formula, the minimum required sample size in each group was 41. After applying prespecified eligibility criteria, diagnostic verification, and data-quality checks, 55 participants met criteria for GDM and 143 for NGT, thereby exceeding the minimum analyzable sample size of 41 per group, resulting in a case–control ratio (∼1:3) that reflects underlying case availability and is appropriate for an unmatched case–control design. (Fig 1)

### Laboratory Analysis

#### Glucose measurement

Blood samples (2 mL) were collected at fasting, 1-hour, and 2-hour time points during the OGTT and analysed on the same day using the glucose-oxidase method (Beckman Coulter, Siemens, Germany). Samples were centrifuged to separate plasma. The coefficient of variation for glucose measurement was 2.02% for low-level and 2.07% for high-level values.

#### Stool IAP measurement

After homogenization, stool supernatant was collected and analyzed for alkaline phosphatase (AP) activity using a standardized protocol on an automated biochemistry analyzer (Nanjing, Jiangsu, China). Briefly, 20 μL of supernatant was added to 1 mL of assay buffer containing 1.25 M diethanolamine (pH 10.2), 0.6 mM magnesium chloride, and 10 mM p-nitrophenyl phosphate as substrate. The reaction mixture was incubated at 37°C for 1 minute, and AP activity was subsequently quantified by colorimetric detection using pre-calibrated AP standards. To assess intestinal alkaline phosphatase specifically, each specimen was additionally treated for 10 minutes using L-phenylalanine (L-Phe, 10 mM final concentration), a specific inhibitor of IAP [17], and then assayed for AP activity using the analyzer. As intestinal alkaline phosphatase accounts for the majority (about 80%) of total stool alkaline phosphatase activity, the stool AP values were expressed as units of IAP/g stool. All measurements were performed by a single technologist who was blinded to participants’ diagnoses, and the assay’s coefficient of variation was 5.06%.

### Statistical Analysis

Data distribution was assessed using the Shapiro-Wilk test. Categorical variables are presented as numbers (%), whereas continuous variables are reported as mean ± standard deviation or median (interquartile range), as appropriate. Comparisons between the GDM and NGT groups were performed using the Student t-test for normally distributed continuous variables and the Mann-Whitney U test for non-normally distributed variables. Categorical variables were compared using the chi-square test. Receiver operating characteristic (ROC) curve analysis was performed to predict the sensitivity and specificity of IAPD in relation to abnormal glucose tolerance during pregnancy. Logistic regression analysis was subsequently performed to identify independent predictors of GDM. Logistic regression variables were selected based on clinical relevance and variables associated with GDM in univariate analysis. Multicollinearity was assessed before model entry. Model fit was evaluated using the Hosmer–Lemeshow test. All tests were two-sided. All statistical analyses were conducted using SPSS version 25.0 (IBM Corp.), and a two-sided P-value of <0.05 was considered statistically significant. Participants with diabetes in pregnancy, inadequate stool samples, or sample quality-control failures were excluded prior to analysis, as shown in Fig 1. Complete-case analysis was used. No variable-level missing data were present in the final analytic dataset. This manuscript was prepared in accordance with the STROBE reporting guideline (S1 Checklist).

## Results

### Baseline characteristics

Table 1 demonstrates the baseline characteristics of the study participants (n = 198). Women with GDM demonstrated significantly higher body weight [62.1 (55.0 – 73.5) vs. 57.5 kg (50.7 – 67.4), p = 0.017] and BMI [median 27.0 (IQR: 23.2 – 31.9) vs. 24.6 (IQR: 21.7 – 28.8) kg/m^2^, p = 0.015] compared to the NGT group. A history of GDM was significantly more common in the current GDM group than in the NGT group (7.3% vs 0.7%, p = 0.022). No significant differences were observed in maternal age, gestational age at enrollment, height, blood pressure, parity distribution, family history of diabetes mellitus, or bad obstetric history parameters between the two groups.

**Table 1.**
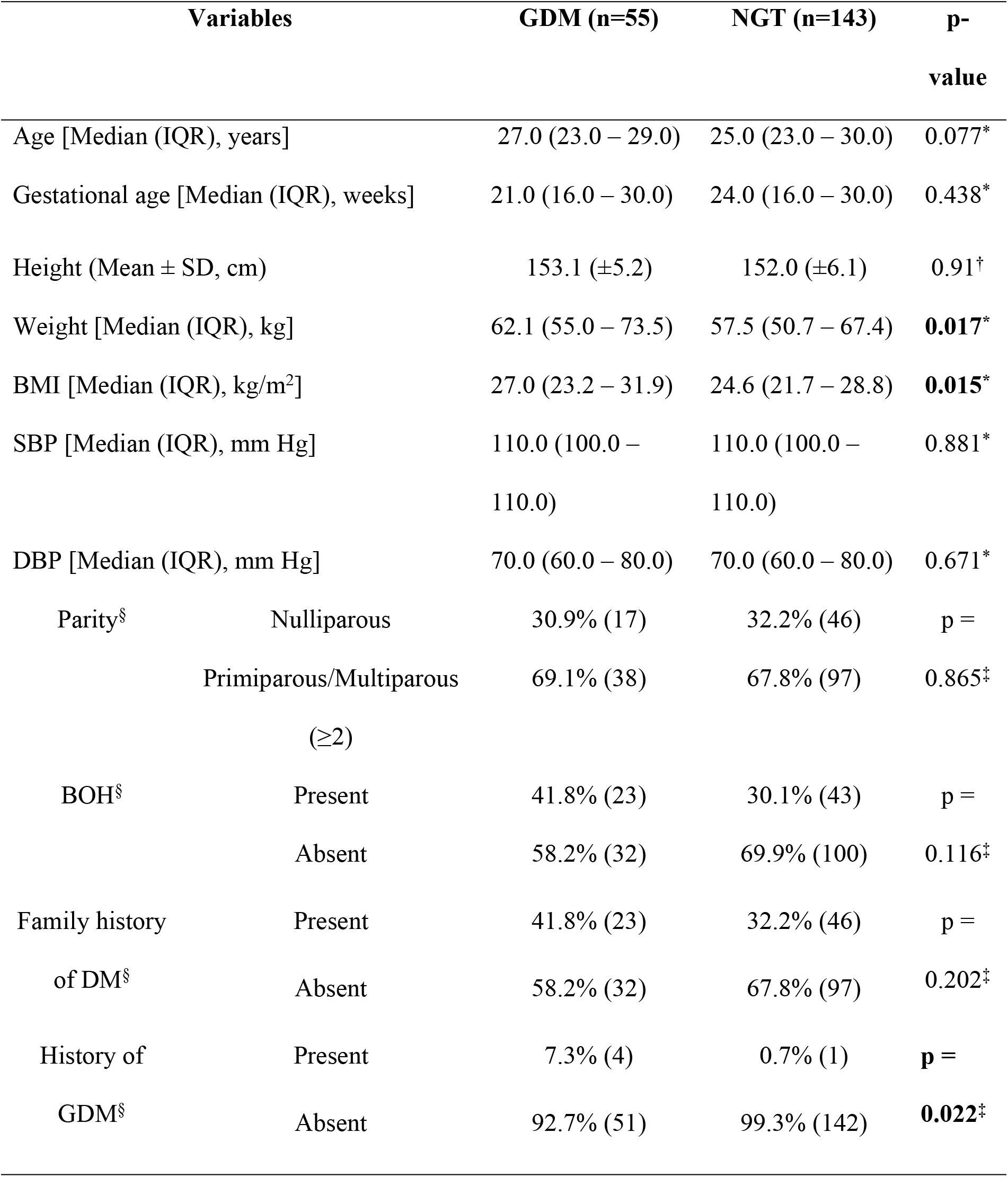
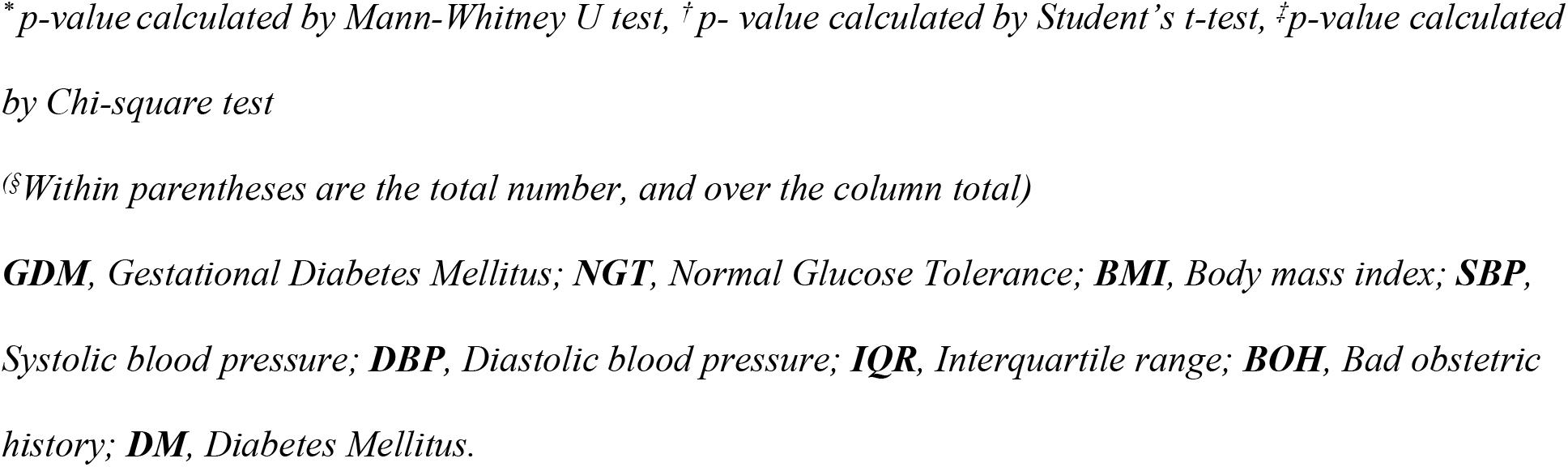
Baseline Characteristics of study participants (n=198)

| Variables |  | GDM (n=55) | NGT (n=143) | p-value |
| --- | --- | --- | --- | --- |
| Age [Median (IQR), years] |  | 27.0 (23.0 – 29.0) | 25.0 (23.0 – 30.0) | 0.077* |
| Gestational age [Median (IQR), weeks] |  | 21.0 (16.0 – 30.0) | 24.0 (16.0 – 30.0) | 0.438* |
| Height (Mean ± SD, cm) |  | 153.1 (±5.2) | 152.0 (±6.1) | 0.91† |
| Weight [Median (IQR), kg] |  | 62.1 (55.0 – 73.5) | 57.5 (50.7 – 67.4) | **0.017\*** |
| BMI [Median (IQR), kg/m<sup>2</sup>] |  | 27.0 (23.2 – 31.9) | 24.6 (21.7 – 28.8) | **0.015\*** |
| SBP [Median (IQR), mm Hg] |  | 110.0 (100.0 – 110.0) | 110.0 (100.0 – 110.0) | 0.881* |
| DBP [Median (IQR), mm Hg] |  | 70.0 (60.0 – 80.0) | 70.0 (60.0 – 80.0) | 0.671* |
| Parity<sup>§</sup> | Nulliparous | 30.9% (17) | 32.2% (46) | p = |
|  | Primiparous/Multiparous (≥2) | 69.1% (38) | 67.8% (97) | 0.865‡ |
| BOH<sup>§</sup> | Present | 41.8% (23) | 30.1% (43) | p = |
|  | Absent | 58.2% (32) | 69.9% (100) | 0.116‡ |
| Family history of DM<sup>§</sup> | Present | 41.8% (23) | 32.2% (46) | p = |
|  | Absent | 58.2% (32) | 67.8% (97) | 0.202‡ |
| History of GDM<sup>§</sup> | Present | 7.3% (4) | 0.7% (1) | **p =** |
|  | Absent | 92.7% (51) | 99.3% (142) | **0.022‡** |

\**p*-value calculated by Mann-Whitney U test, †*p*-value calculated by Student's *t*-test, ‡*p*-value calculated by Chi-square test
§Within parentheses are the total number, and over the column total)
**GDM**, Gestational Diabetes Mellitus; **NGT**, Normal Glucose Tolerance; **BMI**, Body mass index; **SBP**, Systolic blood pressure; **DBP**, Diastolic blood pressure; **IQR**, Interquartile range; **BOH**, Bad obstetric history; **DM**, Diabetes Mellitus.

### IAP level of study participants

In Table 2, IAP levels demonstrated a statistically significant difference between the study groups. Women with GDM had markedly lower median IAP levels than those with NGT (23.59 vs 46.48 U/g of stool; median, p<0.001). The interquartile ranges showed considerable overlap, but lower values were observed in the GDM group (11.95-47.89 U/g) compared to the NGT participants (18.67-104.24 U/g), indicating a substantial IAP deficiency in participants with gestational diabetes. Exploratory IAP deficiency categories based on prior literature, revealing significant differences in distribution between groups (χ^2^ = 16.540, p<0.001). Severe IAPD (0-18 U/g) was more prevalent in GDM participants (43.6% vs 24.5%). Mild deficiency (18.1-65.0 U/g) was also higher in the GDM group (43.6% vs 32.9%). Conversely, normal IAP levels (>65 U/g) were significantly more common in NGT participants (42.7% vs 12.7%), suggesting a stronger association between IAP deficiency and gestational dysglycemia.

**Table 2.**
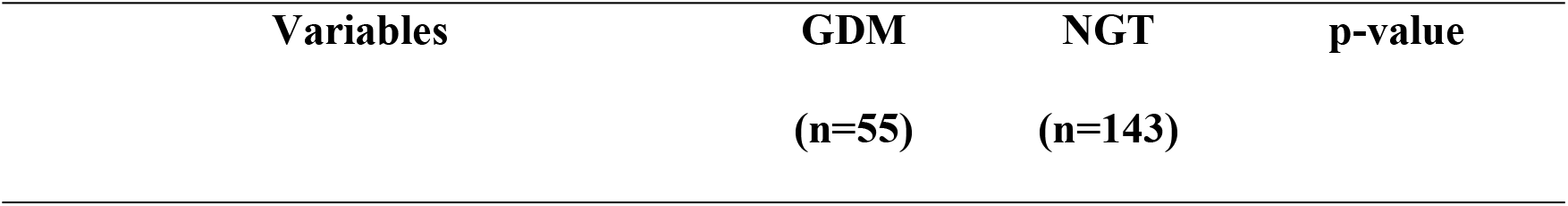
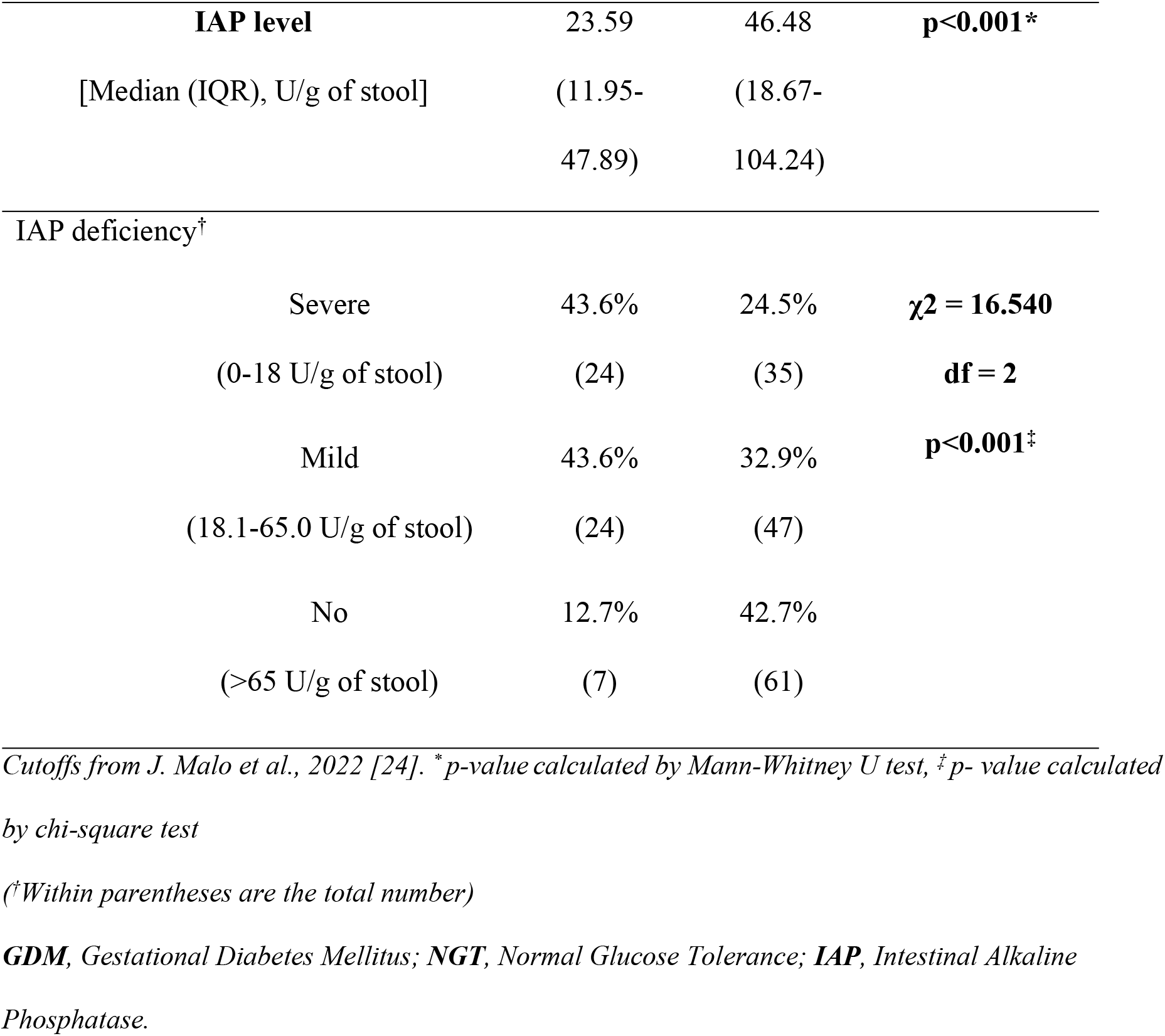
IAP level and grading of IAP deficiency of study participants (n=198)

### Predictors of GDM

Table 3 presents both univariate and multivariate logistic regression analyses to identify significant independent predictors of GDM. In the multivariate model, after adjusting for confounding variables, a lower stool IAP level remained a strong and independent predictor of GDM, with an odds ratio of 0.98 (95% CI 0.97 to 0.99; p<0.001). This indicates that for every one-unit increase in stool IAP, the odds of having GDM decrease by 2%.

**Table 3.** Univariate and multivariate logistic regression analysis of predictors associated with the development of GDM.

| Variables | Univariate |  | Multivariate |  |
| --- | --- | --- | --- | --- |
|  | Odds Ratio (95% CI) | p-value | Odds Ratio (95% CI) | p-value* |
| Age | **1.06 (1.0 – 1.1)** | **0.05** | 1.04 (0.97 – 1.11) | 0.296 |
| Gestational age | 0.985 (0.949 – 1.021) | 0.405 | - | - |
| IAP | **0.98 (0.98 – 0.99)** | **<0.001** | **0.98 (0.97 – 0.99)** | **<0.001** |
| F/H/O DM | 1.52 (0.8 – 2.88) | 0.203 | - | - |
| P/H/O GDM | **11.14 (1.22 – 101.99)** | **0.033** | **10.05 (1.06 – 95.53)** | **0.045** |
| BMI (kg/m<sup>2</sup>) | **1.07 (1.01 – 1.14)** | **0.027** | **1.08 (1 – 1.15)** | **0.032** |

\*Nagelkerke *R* square: 0.22, \*Hosmer & Lemeshow test *p* = 0.748
**GDM**, Gestational Diabetes Mellitus; **IAP**, Intestinal Alkaline Phosphatase; **F/H/O DM**, Family history of Diabetes Mellitus; **P/H/O GDM**, Previous history of Gestational Diabetes Mellitus; **BMI**, Body mass index.

### IAP as a predictor of GDM

Fig 2 illustrates the ROC curve for predicting IAP as a predictor of GDM. Here, the area under the curve is 0.676, 95% CI: 0.599 – 0.753, p-value < 0.001. The Youden index (sensitivity + specificity - 1) was calculated at different reference points. At a stool IAP threshold of 65.4 U/g, sensitivity for identifying GDM status was 89.1%, with a specificity of 42.0%.

**Fig 2.**
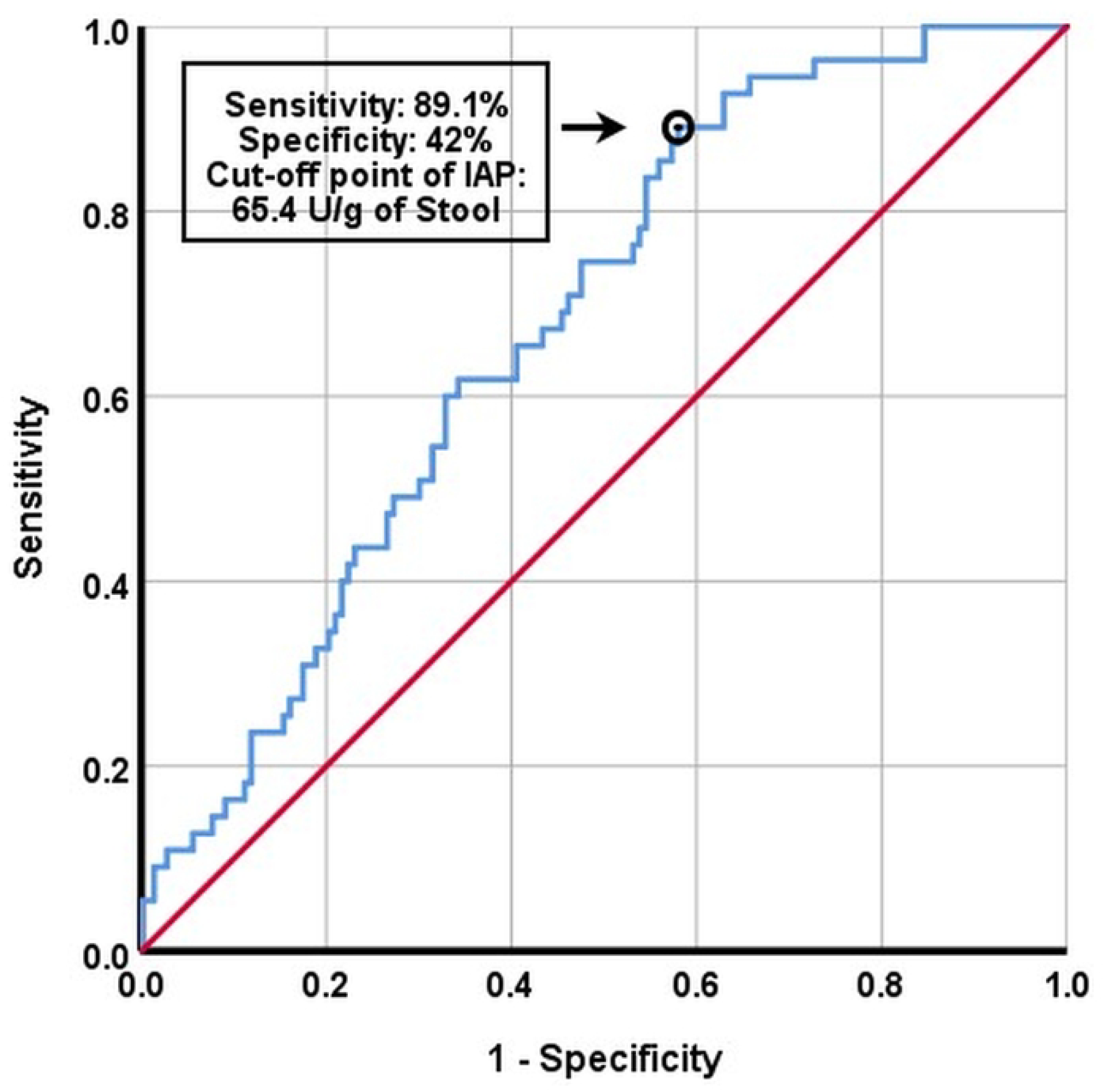
ROC curve for IAP as a predictor of GDM. The area under the curve is 0.676, 95% CI: 0.599 – 0.753, p-value < 0.001. The Youden index (sensitivity + specificity - 1) was calculated at different reference points. At a stool IAP threshold of 65.4 U/g (the highest value), sensitivity for identifying GDM status was 89.1%, with specificity of 42.0%. Abbreviations: **GDM**, Gestational Diabetes Mellitus; **IAP**, Intestinal Alkaline Phosphatase; **ROC**, Receiver operating characteristic.

## Discussion

The principal finding of this study is that fecal IAP levels are markedly reduced in women with GDM compared with those of normoglycemic pregnant women. Furthermore, a lower IAP level remained independently associated with GDM after adjustment for established clinical risk factors and the observed graded association between declining IAP level and the presence of GDM, suggesting that it may represent an under-recognized component of the metabolic disturbances associated with gestational dysglycemia. The importance of these findings lies in their novelty and potential translational implications. To our knowledge, this study represents the first investigation to evaluate fecal IAP levels in relation to gestational dysglycemia. Although stool IAP alone does not demonstrate sufficient discriminative performance to serve as a standalone diagnostic marker, its high sensitivity suggests it may have utility as an adjunct biomarker for identifying women at increased risk of GDM.

This study has several strengths. It represents the first evaluation of fecal IAP level in the context of gestational diabetes, thereby addressing an important gap in the literature. The use of standardized laboratory measurements and adjustment for key clinical confounders enhances the robustness of the observed associations. Furthermore, the identification of a gut-derived biomarker contributes to the growing recognition of the gut–metabolic axis as an important contributor to metabolic disorders during pregnancy. Several limitations should also be acknowledged. The cross-sectional design limits the ability to determine temporal relationships. Residual confounding from unmeasured factors such as diet and microbiome composition cannot be excluded. In addition, the study was conducted in three antenatal care clinics across a single region of Bangladesh, which may limit generalizability to populations with different microbiota compositions or genetic backgrounds. As this was a hospital-based study, selection bias cannot be excluded. Finally, the study did not include measurements of related microbiome markers, which could have provided further mechanistic insight into the gut-immune pathways.

Baseline demographic and obstetric characteristics were broadly similar between groups; however, body weight and BMI were higher in women with GDM, consistent with the established role of obesity in insulin resistance during pregnancy [25]. A prior history of GDM was also more common in the GDM group, in keeping with its known recurrence risk [26]. The findings of a marked reduction in stool IAP levels in GDM compared with those in NGT are consistent with those from non-pregnant populations, which demonstrate lower IAP levels in individuals with type 2 diabetes mellitus (T2DM), and suggest a protective association of higher IAP with dysglycaemia [18]. As GDM and T2DM share a common pathophysiological pathway, comprising β-cell dysfunction and insulin resistance, this alignment strengthens the biological plausibility that IAP deficiency may reflect a shared metabolic pathway linking intestinal dysfunction and dysglycemia [25,27].

The findings may have important explanatory and clinical implications. Mechanistically, IAP helps maintain intestinal barrier integrity and regulate host–microbiome interactions by detoxifying lipopolysaccharides and limiting intestinal permeability, metabolic endotoxemia, and downstream inflammatory signaling [15–17]. Reduced IAP has been linked to low-grade inflammation and impaired metabolic regulation in both experimental and clinical studies. Because pregnancy is already a physiologically insulin-resistant state, IAP deficiency may further amplify inflammatory and metabolic disturbances that predispose susceptible women to gestational dysglycemia. When IAP level was examined across deficiency categories, a gradient in GDM frequency was observed, suggesting an exposure–response relationship between declining IAP level and metabolic disturbance. Multivariable analyses further demonstrated that a reduced stool IAP level remained independently associated with GDM, even after adjustment for important confounders, including body mass index and prior GDM. This finding suggests that IAP deficiency may reflect a component of gut-metabolic dysregulation, rather than merely reflecting established clinical risk factors. Interestingly, the optimal threshold identified in this study was comparable to values reported in non-pregnant populations. This observation suggests that the physiological hormonal changes of pregnancy, including placental hormone secretion, may not substantially alter the relationship between IAP level and metabolic regulation. Rather, reduced IAP appears to reflect underlying metabolic disturbances related to glucose rather than pregnancy-specific physiological adaptations. Nevertheless, the modest predictive performance observed indicates that IAP should not be considered a replacement for established diagnostic tests. Instead, it may represent one component within a broader network of metabolic, inflammatory, and microbiome-related pathways involved in the pathogenesis of gestational glucose intolerance.

Several unanswered questions remain. A substantial proportion of normoglycemic participants also demonstrated varying degrees of IAP deficiency. These individuals may represent a subgroup with increased metabolic susceptibility who could be at elevated risk of developing gestational diabetes in future pregnancies or metabolic disorders later in life. This observation aligns with the concept of “incipient metabolic syndrome” described in relation to fecal IAP deficiency and with longitudinal evidence linking persistent IAP deficiency with an increased risk of developing diabetes in non-pregnant populations in later life [18,24]. This subgroup may warrant prospective follow-up.

## Conclusion

This study provides novel evidence that reduced fecal IAP level is independently associated with GDM. These findings extend current understanding of gestational diabetes beyond traditional established endocrine pathways, highlighting the potential contribution of gut-derived metabolic regulation. While stool IAP measurement cannot replace the oral glucose tolerance test as a diagnostic tool, it may hold potential as a complementary biomarker for metabolic risk stratification during pregnancy. Prospective studies integrating microbiome, inflammatory, and metabolic profiling are needed to clarify its predictive and preventive relevance.

## Data Availability

The dataset generated and/or analyzed during the current study is not publicly available because it contains potentially sensitive information, and public sharing could compromise participant confidentiality. De-identified data relevant to the findings of this study may be made available to qualified researchers upon reasonable request and subject to approval by the Institutional Review Board of Bangladesh Medical University, Shahbagh, Dhaka. Requests for data access should be directed to the Institutional Review Board at, referencing study registration number 4823 (BSMMU/2024/2012 approved on 25 February 2024). Data will be shared in accordance with participant consent, institutional policies, and applicable ethical and legal requirements.

## Acknowledgments

We extend our sincere gratitude to all the participants of the study. Special thanks to the Gestational Diabetes Mellitus (GDM) group members for their efforts in data collection. We also acknowledge the technical contributions of the laboratory staff at the Stap BioTech lab in Ashulia, Dhaka, to the rapid transport of stool samples and timely processing.

## Supporting information

S1 Checklist. STROBE Checklist

